# Towards long term monitoring: Seizure detection with reduced electroencephalogram channels

**DOI:** 10.1101/2021.12.14.21267701

**Authors:** Christina Maher, Yikai Yang, Nhan Duy Truong, Chenyu Wang, Armin Nikpour, Omid Kavehei

## Abstract

Epilepsy is a prevalent condition characterised by recurrent, unpredictable seizures. The diagnosis of epilepsy is by surface electroencephalography (EEG), a time-consuming and uncomfortable process for patients. The diagnosis of seizures using EEG over a brief monitoring period has variable success, dependent on patient tolerance and seizure frequency. Further, the availability of hospital resources, and hardware and software specifications inherently limit the capacity to perform long-term data collection whilst maintaining patient comfort. The application and maintenance of the standard number of electrodes restrict recording time to a maximum of approximately ten days. This limited monitoring period also results in limited data for machine learning models for seizure detection and classification. This work examines the literature on the impact of reduced electrodes on data accuracy and reliability in seizure detection. We present two electrode ranking models that demonstrate the decline in seizure detection performance associated with reducing electrodes. We assert the need for further research in electrode reduction to advance solutions toward portable, reliable devices that can simultaneously provide patient comfort, long-term monitoring and contribute to multimodal patient care solutions.

## 1. Introduction

Epilepsy is a severe neurological condition that affects millions of people world-wide. The main symptoms are recurrent seizures that can be a traumatic experience for the individual [1]. Epilepsy diagnosis requires attendance at a specialised epilepsy clinic or hospital. The lengthy process involves multiple tests, continuous electroencephalography (EEG) and video EEG (vEEG) monitoring, and if no seizures are recorded, may result in misdiagnosis. Currently, this process is conducted either in a hospital ward or via an ambulatory service by wearing the EEG device at home for several days. Additionally, a carer is usually required to be present for ward testing.

EEG data analysis is a lengthy process, mainly if seizures are not immediately evident on the surface EEG or vEEG. There may be no or little surface EEG change during focal seizures without impaired awareness. The incidence of false-positive diagnosis reportedly ranges from 2%-71% [2]. EEG monitoring is costly in financial, mental, physical and time resources. In 2016, the global prevalence of active epilepsy was 45.9 million individuals with 126, 055 epilepsy-related deaths, 13.5 million disability-adjusted-life-years, 5.9 million years of life lost (YLL), and 7.5 million years of life with a disability (YLD) [3]. The risk of misdiagnosis must be weighed against that of a false positive diagnosis [2]. Therefore, improving EEG device portability and automated detection represents an essential advancement to facilitate effective diagnosis and treatment and enhance the patient journey.

This work serves to explain the routine clinical seizure detection process and present two channel selection algorithms developed in our lab. Our channel selection algorithms were applied to publicly available datasets, highlighting the difficulty in achieving high seizure detection performance with a lower number of channels. We discuss previous work that attempted to reduce the number of electrodes used in automated seizure detection models, and suggest their applications for improving long term monitoring solutions.

### 1.1. The need for electrode reduction

Several factors support the focus on reducing the number of electrodes used in seizure detection. Technological advances have made automated seizure detection and device portability a near reality. Currently, the longest non-invasive surface EEG recording using wet or dry electrodes is around ten days. Applying abrasive gel to the scalp can irritate and damage the skin, yet it is compulsory for standard EEG electrodes. Mobile EEG systems must be comfortable and appealing to wear. Thus the portability of EEG systems represents a trade-off between electrode count and the quality of the signal. Achieving this balance in a portable EEG system would enable home EEG monitoring, potentially greatly improving patient comfort during the diagnosis process. Fig. 1 depicts the patient journey through the healthcare system from the first seizure through to epilepsy diagnosis.

**Figure 1.**
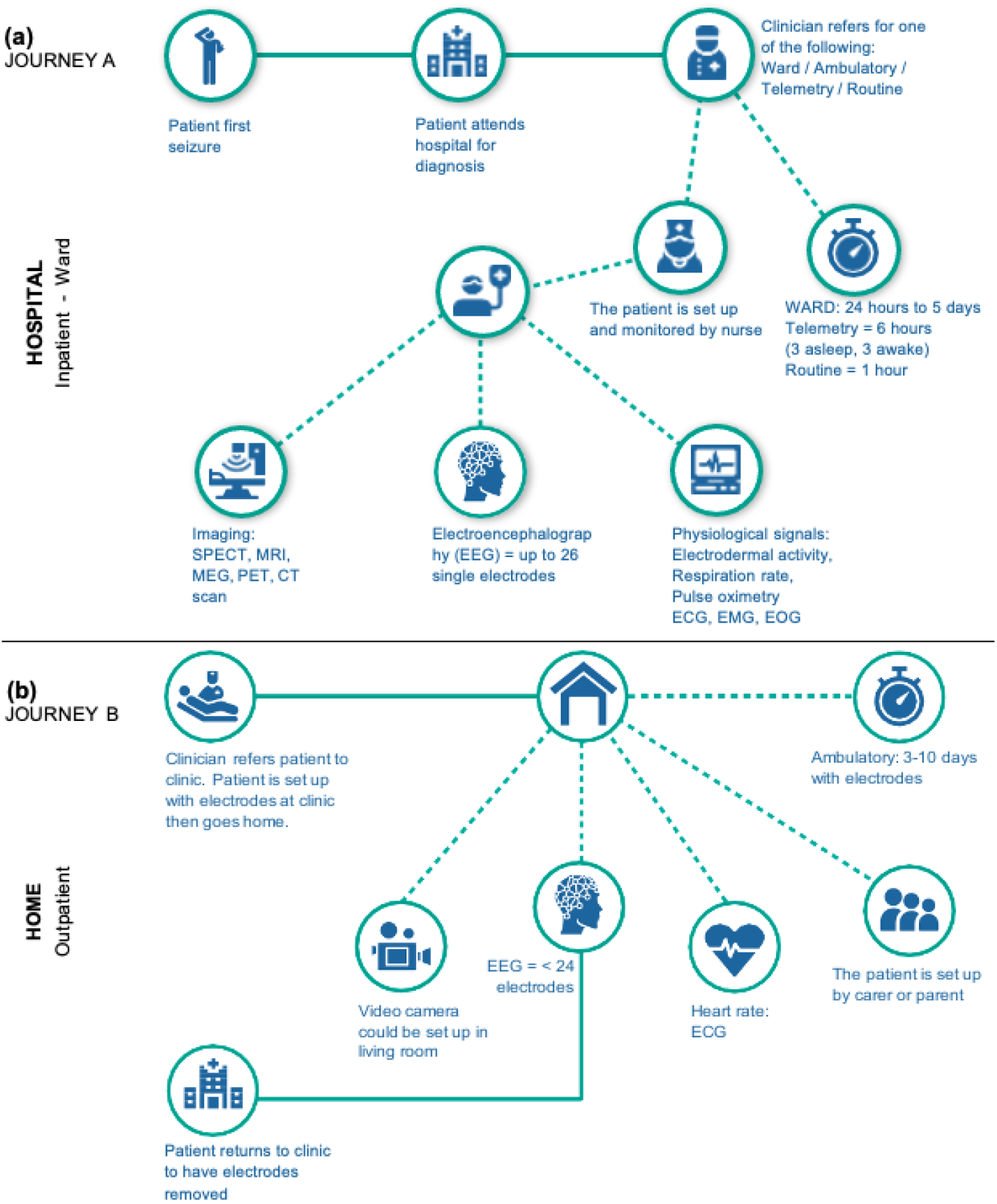
Patient journey - two scenarios. This figure depicts the process and various settings where seizure detection and logging may take place. In the hospital setting (a, Journey A), a carer accompanies the patient. Patient comfort, mobility, and limited hospital resources restrict the length of time a patient can spend in the hospital. Conversely, the home setting (b, Journey B) facilitates patient comfort and mobility. Patients can continue their daily activities while being monitored for an extended duration. Unfortunately, improving patient comfort and mobility through home monitoring comes at the cost of diminished EEG recordings due to limitations imposed by the individual and their environment. For example, electrodes do not last long on the scalp, and the patient must inevitably return to the hospital or have a technician visit them to adjust or remove the electrodes. Avenues for improving this imperfect process lie in advancing the research in electrode reduction to enable long-term wearable solutions for the patient.

Herta [4] and colleagues emphasised the value of fewer electrodes in circumstances where a 10-20 EEG system cannot be applied, such as in intensive care units (ICUs). Ambulatory monitoring is a potential beneficiary of reduced electrodes through automated seizure logging, which is currently achieved through either ward monitoring or self-report. However, the accuracy of self-report is highly dependent on seizure-type, and patient ability [34]. Inaccurate seizure reporting impacts patient treatment and conflates reporting on the efficacy of treatments such as medication. Under-reporting of seizures is a long-standing and significant clinical problem [34–42]. Sub scalp electrodes, if shown to be a reliable method for recording EEG, can replace the need for patients to regularly attend clinics to have electrodes re-applied. Moreover, subcutaneous electrode systems purport to have a high agreement with surface EEG recordings [5]. Yet the number of electrodes in subcutaneous systems remains restricted; more than four electrodes are not ideal. Hence, improving automated seizure detection performance using a reduced number of surface electrodes is the first step towards reliable, long-term EEG monitoring solutions that can improve the patient journey.

Furthermore, patient-specific electrode reduction would improve recording of their specific seizure semiology. The number of required electrodes and electrode placement can be guided by a patient’s magnetic resonance imaging (MRI) scan and EEG. We recently examined the role of structural connectivity in seizure propagation in focal epilepsy patients [6]. Diffusion MRI can be a valuable aid for patient-specific determination of electrode placement. This paper is intended to extend our previous work by examining the best automated method for electrode based on EEG signals. Taken together, the individualised MRI data combined with the automated ranking of EEG signals can provide a patient-specific pipeline for seizure onset zone selection, which would indicate the optimal electrode placement and the number of electrodes to optimise automated seizure detection. Fig. 2 portrays a theoretical process for using both EEG and MRI to obtain the optimal electrode placement. Items (a) and (b) were explored in our previous work [6] whilst (c) and (d), circled in red, are examined in Section 4 of this paper.

**Figure 2.**
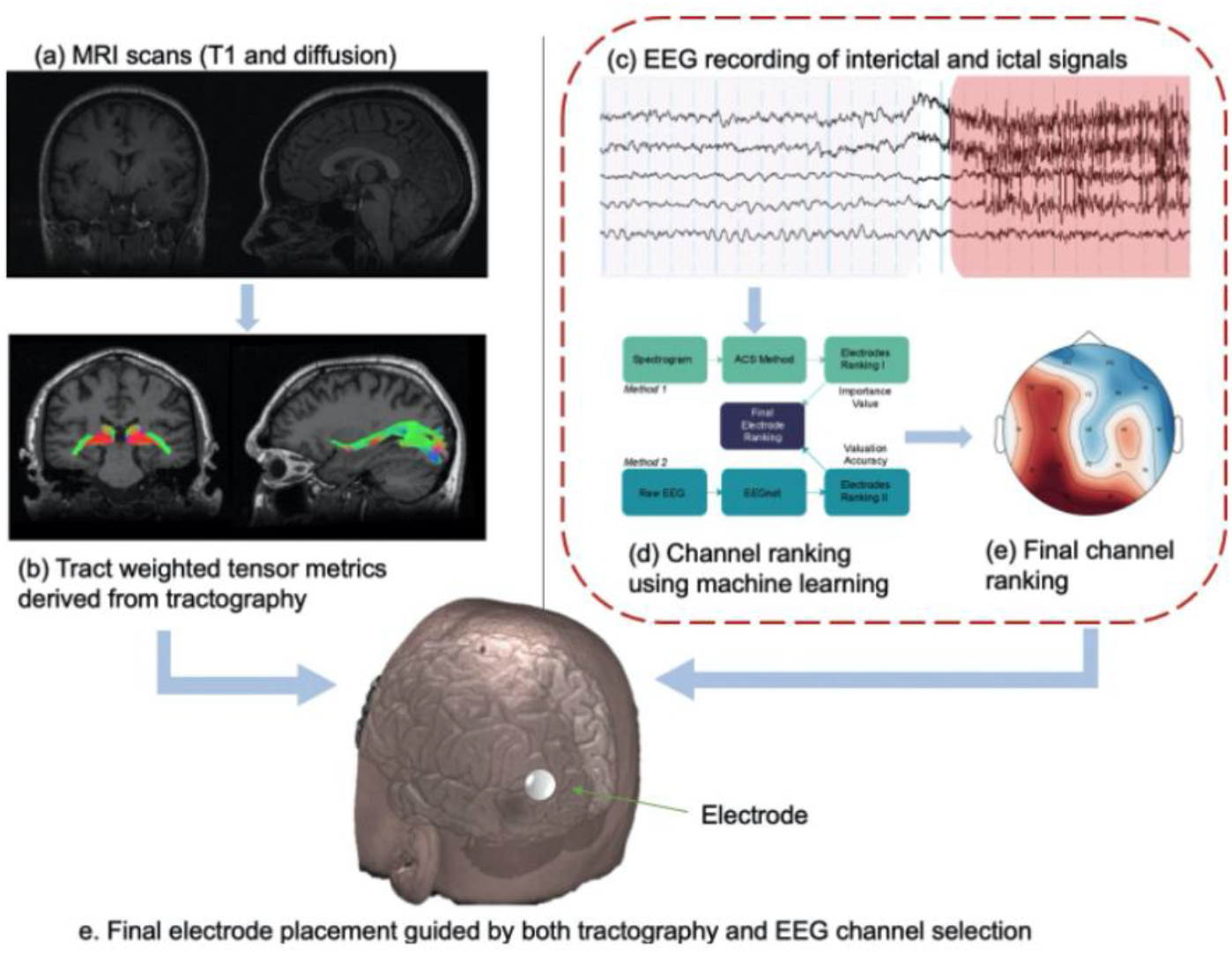
MRI and EEG for optimal electrode placement. The MRI scans (T1 and diffusion) (a) are processed through an imaging pipeline to obtain tractography and tract-weighted tensor metrics (b). The EEG signals (c) are read and seizures detected using machine learning (ML) (d) and the most important electrodes per patient are selected (e). Lastly, the output of both the tract-weighted tensor metrics and the EEG electrode selection can be used by the clinicians to select the optimal placement for a wearable or implantable electrode.

## 2. Manual seizure detection

Seizures are detected by monitoring the EEG signals. EEG is the gold standard used to detect interictal, ictal and subclinical epileptic activity. This includes critical conditions like status epilepticus [7,8]. Changes in EEG signals could represent spontaneous electrical brain activities that exhibit dynamic, stochastic, nonlinear, non-stationary, and complex behaviour with high temporal resolution. Typical EEG monitoring requires up to 19 head, and two reference electrodes, based on the international 10–20 electrode placement system [9]. This standard electrode placement is applied in a variety of settings, including for inpatients in the hospital. In the context of ambulatory monitoring in the home setting, a device with up to 19 electrodes may be used. For seizure monitoring in neonates in the ICU, four electrodes are considered sufficient. The routine practice of continuous EEG monitoring in ICUs affords a consensus on the reliability of a reduced number of electrodes [10].

Electrode positioning and a typical ictal event are demonstrated in Fig. 3. Here, an interictal period precedes an ictal event (red highlighted section), shown on a montage of five channels out of the standard 21. In routine clinical practice, seizure onset and type are identified, labelled and classified. Human experts (“expert”) may be nurses, EEG technicians, neurophysiologists and neurologists, trained to detect seizures on the EEG. The expert then labels the EEG with the seizure time, length and location. Usually, EEG data recorded over an extended period yields reliable, interpretable EEG that can inform clinical decisions.

**Figure 3.**
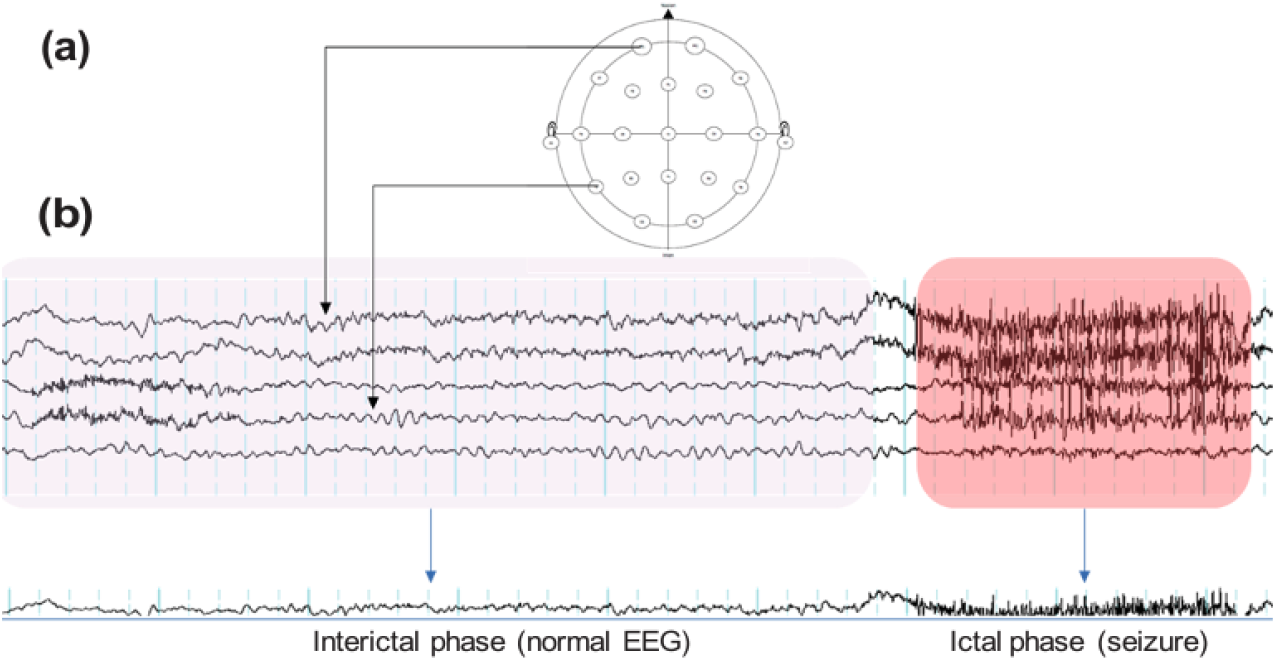
EEG signals are recorded from electrodes placed on the scalp in the 10-20 format (a). The signals are then displayed on readout software. An EEG technician or neurologist then visually identifies and labels the interictal, pre-ictal and ictal phases (b).

## 3. Automated seizure detection

Computer-automated techniques, such as ML algorithms, can detect and label seizures. EEG signal recordings and the amount of EEG recorded play a fundamental role in the outcome of the seizure detection model [11]. Channel selection involves reducing the number of electrodes to be analysed or utilising a range of feature selection and classification techniques, or a combination of both [44,45,49,60]. The model is trained either on individual patients or across all patients and then tested on the dataset of interest [12,13].

Digital signal processing allows the extraction of common features of an EEG signal. Prior human studies examine engineered features such as relative average amplitude, relative scale energy, average cross-correlation function, relative power, bounded variation [14], phase-amplitude coupling [15], root mean square [16], energy [17], median frequency [18], entropy [19,20], correlation dimension [21,22], maximal Lyapunov exponent [23], and skewness and kurtosis [24]. These features may be the sole focus of the model [20] or combined and applied to a selected EEG frequency band [25–27]. There are also options for direct extraction of features from raw EEG, or after a filter such as short-time Fourier transform (STFT) or wavelet transform is applied to the EEG.

In conventional ML methods, such as convolutional neural networks (CNN), the aim is to classify EEG signals [27,28]. EEG features are fed into the model, and additional threshold mechanisms are then applied to the output of the training model via a “classifier” technique [29]. Common classifier techniques include but are not limited to Bayesian, K-nearest neighbours (KNN), decision tree, random forest, and support vector machine (SVM) classifiers [11]. Biomarkers for seizure susceptibility may help improve automated seizure detection models, yet tools for such exploration are limited. Our recent work examined biomarkers for seizure susceptibility using a Bayesian CNN based tool [31]. We showed interictal slowing activity was a promising feature for the investigation of seizure susceptibility prediction.

The reliance of automated seizure detection models on engineered features emphasises the vulnerability of the model to changes in the electrode count, EEG artefacts and noise [30]. Individual human characteristics, such as hair and scalp thickness, and pathological state, also influence EEG recordings. Reducing electrodes whilst maintaining the fidelity of recordings demands the ML model and the electrode placement be patient-specific. Therefore electrode reduction is currently considered problematic and receives little enthusiasm in the clinical setting. The increased availability of sufficient and reliable EEG data is critical to the acceptance and translation of automated seizure detection into the clinical realm. The need for increased data can be met through a reduced electrode, long-term, implantable device.

## 4. Methods

### 4.1. Channel Selection Method 1

Our group previously used this random forest classifier method on the Kaggle iEEG dataset to rank channels in order of their contribution to the detection of a given seizure [50]. Here we apply it to the EPILEPSIAE scalp EEG dataset to test its utility on a different dataset. The descriptive statistics of the EPILEPSIAE dataset are shown in Fig. 4. The channel selection was patient-specific, meaning the channels selected as the best channels differed from patient to patient. The pseudo-code for the automatic channel selection (ACS) algorithm is shown below.

**Figure 4.**
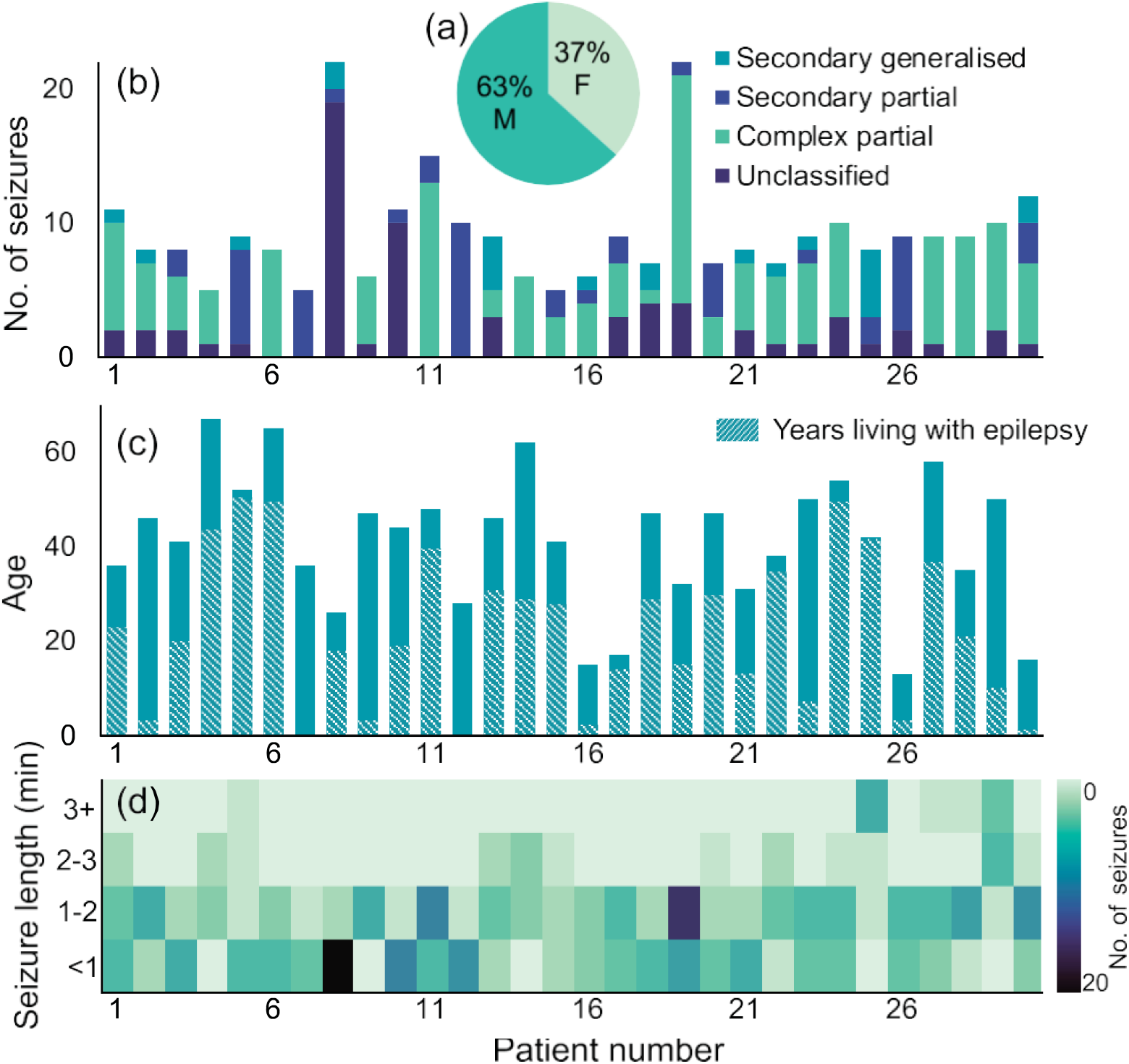
Descriptive statistics from the EPILEPSIAE dataset [51,52]. Gender is shown in (a), seizure type, and the number of seizures per patient is shown in (b). Age distribution (c) includes a shaded portion, which represents the number of years living with epilepsy. The seizure length and frequency of that seizure length is depicted in the heat-map in (c). Seizure frequency is shown in the colour bar, where darker colours represent a higher number of seizures and lighter colours represent a lower number of seizures.

For the current paper, the ACS algorithm was used to select the 10 most important channels per patient on the EPILEPSIAE surface EEG dataset (on 30 patients). Next, the top 10 channels across the entire group of 30 patients were selected and fed into our seizure detection model. To test the performance of the 6 channels, we chose a subset of the top 6 channels from the top 10. The seizure detection model was then applied to the 6 channels. The average AUC score for 19, 10, and 6 channels was 98.03, 91.57, 92.25, respectively. With just 6 channels, we observed an encouraging trend of improved performance on some patients; yet the number of false positives remained a challenge.

### 4.2. Channel Selection Method 2

For this method of channel selection, we performed seizure detection on a subset of the TUH dataset. Descriptive statistics for the TUH dataset are shown in Fig. 5.

**Figure 5.**
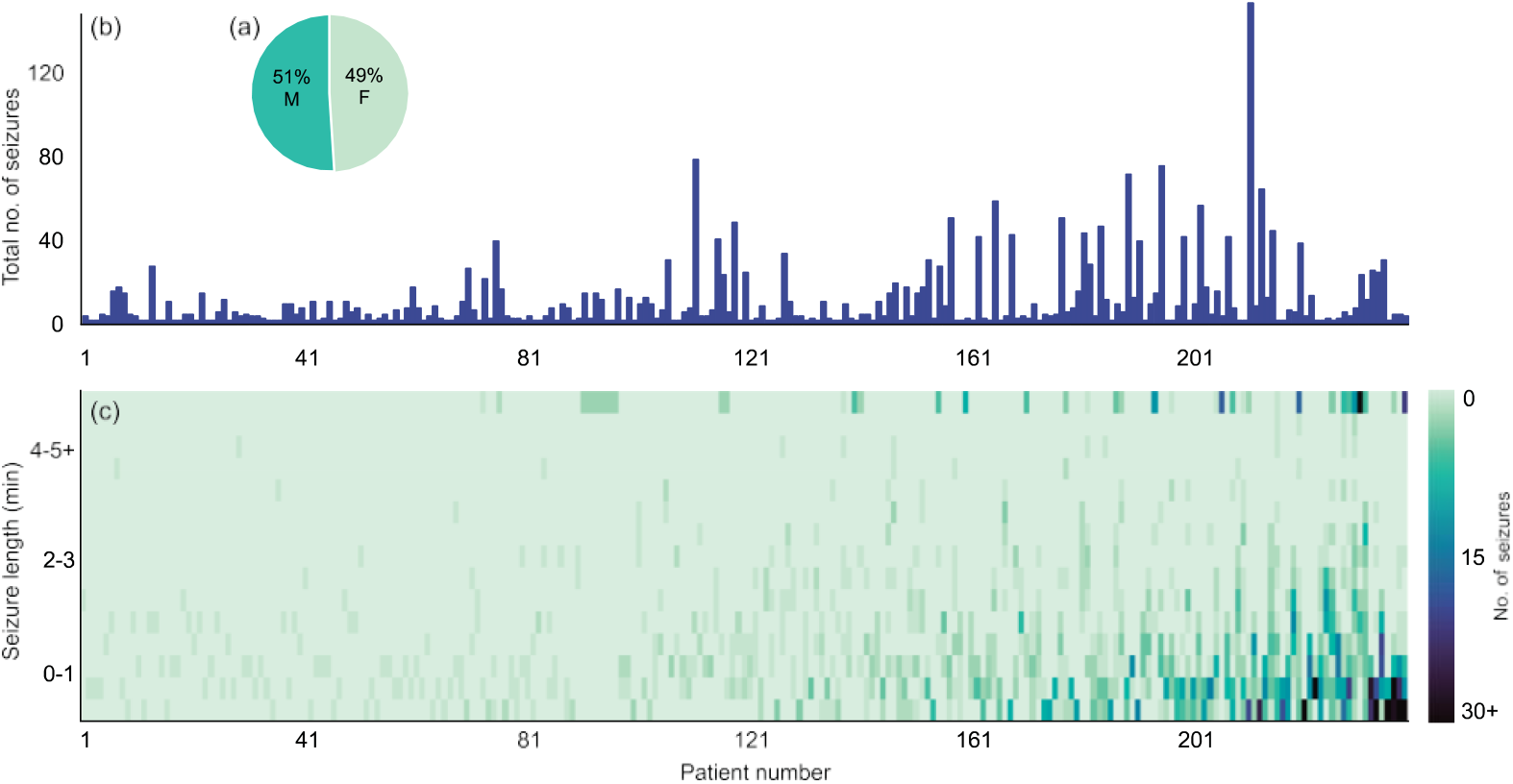
Descriptive statistics from the TUH dataset. Gender distribution is shown in (a), and total number of seizures per patient is shown in (b). The seizure length and frequency of a specific seizure length is depicted in the heat-map in (c). Seizure frequency is shown in the colour bar, where darker colours represent higher number of seizures and lighter colours represent a lower number of seizures.

Channel selection for this method involves a combination of two approaches:

#### Approach 1

Information from a single electrode was fed into a simple and fast CNN structure (EEGNet, as seen in [53]), which has been widely used in the brain-computer interface domain. Channels were then selected by ranking valuation accuracy. The final channel selection decision was based on the ranking results of the two channels. A further experiment on the high-ranking channels was also conducted (Approach 2). The procedures are shown in Fig. 6.

**Figure 6.**
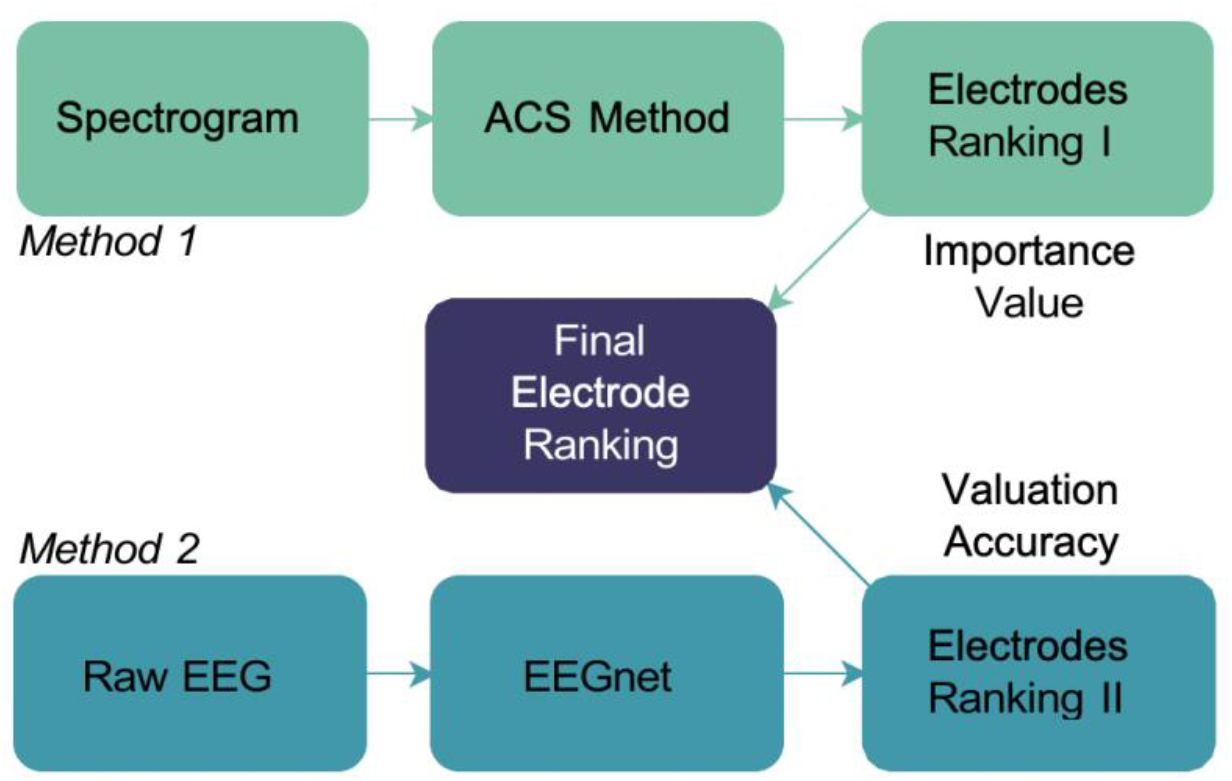
Channel ranking and selection procedure.

#### Approach 2

A fast single electrode training was conducted to rank the channels. The classifier was trained to feed one electrode one time and rank channels based on the valuation accuracy. To make this method less time-costly, we trained only one epoch for comparison as there were a total of 21 electrodes to test. Unlike previously mentioned methods which use frequency information in training, this approach trains the raw EEG directly. The final channel ranking heat-map is shown in Fig. 7.

**Figure 7.**
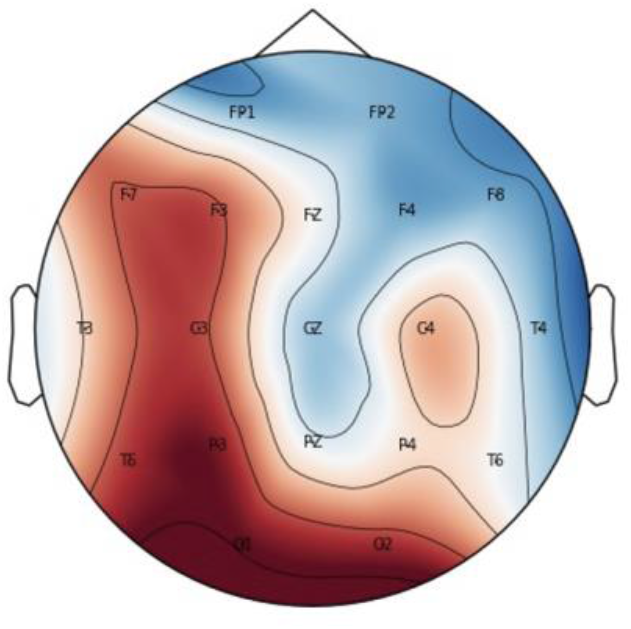
Channel ranking heat-map. Color changing from blue to red indicates increasing ranking of the electrodes.

We combined the results of the two electrode ranking methods based on (1) The importance value from approach one, which reflects how often it is used during training in the random forest classifier; (2) The valuation accuracy from approach two, which reflects the first epoch results when we put a signal electrode into a fast structure training.

##### Algorithm 1 Automatic channel selection

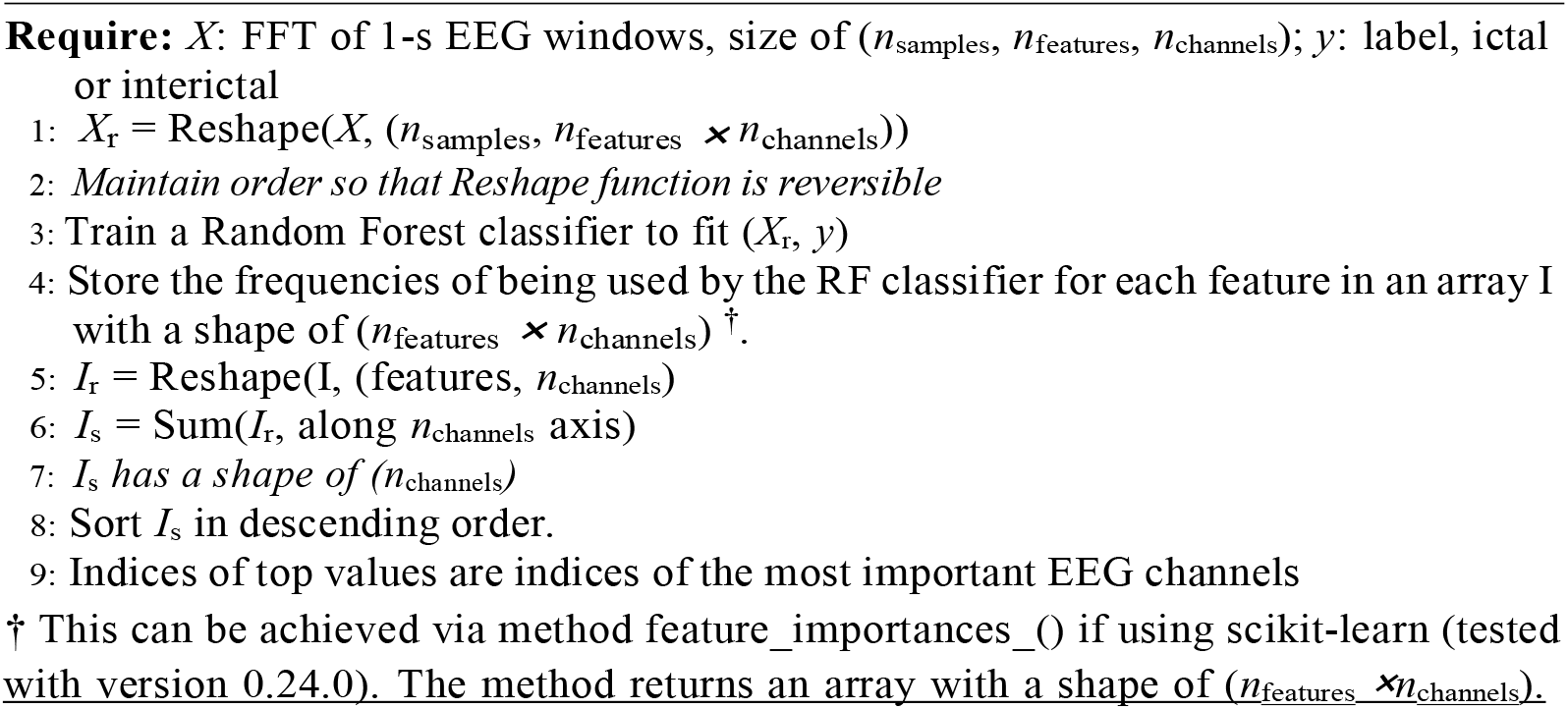

## 5. Results

As seen in Fig. 7, the five electrodes F3, F7, P3, O1, and O2 ranked the highest. The electrodes were paired in the format F3-F7 and P3-O1 as these were two pairs of nearby electrodes, to give the two-channel format that was fed into the model. The purpose of the two approaches for channel selection was to make the selection more reliable. Interestingly, literature shows that left temporal lobe epilepsy is a common epilepsy type, with focal seizures in the left temporal lobe being the most common seizure type. Our model achieved a sensitivity score of 2.04, which reaches 0.17 FA/24 hrs using only two channels. The modest results highlight the challenges in EEG seizure detection with a minimal number of electrodes.

## 6. Discussion

Given the challenges in automated seizure detection and logging, a portable, reduced electrode device would be invaluable for patients. Such a device could materialise as a sub scalp implant with as little as three electrodes tailored to patient-specific seizure detection. A sub scalp implant would address the long-term monitoring needs of individuals with intractable epilepsy who are unsuitable for surgery. The device could act as a monitor to guard against sudden unexpected death in epilepsy (SUDEP), prevalent in low resource countries [54,55]. However, the reduced electrode device must record EEG signals and detect and log seizures reliably to be clinically useful. Unfortunately, research into incumbent devices has yielded mixed results [56]. A summary of prior research is in Table 1.

**Table 1:** Seizure detection with reduced channels, channel selection models, and/or reduced electrodes.

| Reference | Dataset | No. E/C § | Patient specific | No. patients | Age group £ | Total no. seizures | Sensitivity | Specificity |
| --- | --- | --- | --- | --- | --- | --- | --- | --- |
| Birjandtalab *et al.* 2017 [43] | CHB-MIT | 18 E | Y | 23 | P | – | 80.87%(avg) | 47.45% |
| Selvakumari *et al.* 2019 [44] | CHB-MIT | 12 E | Y | 23 | P | – | 95.70% | 96.55% |
| Pisano *et al.* 2020 [45] | EPILEPSIAE (surface) | 11 C | N | 3 | A | 28 | 94% | 95% |
| Herta *et al.* 2017 [4] | General Hospital Vienna † | 6 E | N | 83 | – | – | 84.99–93.39% | 90.05–95.6% |
| Bhattacharyya *et al.* 2017 [46] | CHB-MIT | 5 C | N | 23 | P | – | 97.91% | 99.57% |
| Moctezuma *et al.* 2020 [47] | CHB-MIT | 4-6 C | Y | 23 | P | – | – | – |
| Shah *et al.* 2017 [48] | TUH EEG Seizure Corpus (TUSZ) | 4-2 C | N | – | – | – | – | – |
| Kjaer *et al.* 2017 [49] | Rigshospitalet and Northzealand Hospital ‡ | 3 E | N | 6 | P | ~8.75/pat. | 98.40% | 100% |
| Maher *et al.* 2021 (current paper) | EPILEPSIAE (surface) | 6 C | Y | 30 | A | – | 92.25(AUC) |  |
|  | TUH | 5 C | Y | 249 | A | – | 2.04% |  |

£ A for adult; P for pediatric.
§ E for electrodes; C for channels.
‡ This dataset is from Rigshospitalet and Northzealand Hospital.
† This group reported 9 electrodes had the highest sensitivity, but 6 electrodes performed best on their “RDA” model.

A foundational study by Shih *et al*. used a greedy backward elimination algorithm to select a subset of seven features that produce the lowest false positive rate from each channel [61]. The reduced number of channels was between 18 and 4.6, achieving improvement in false-positive rate (FPR), from 0.35/hr to 0.19/hr, yet sensitivity and detection delay worsened, from 99% to 97% and 7.8 s to 11.2 s, respectively. More recently, Moctezuma and Molinas compared the popular backward greedy elimination algorithm with two versions of the nondominated sorting genetic algorithm (NSGA), NSGA-II and NSGA-III [47]. They achieved an accuracy ranging between 0.98 and 1 using one to two channels, comparable to their detection result with the full electrode count (accuracy ranged from 1 to 0.97).

The role of variance contributed by individual channels has been widely explored. Duun-Henriksen *et al*. reduced channels by selecting the largest variance in channels, achieving a detection performance on three channels near equivalent to a clinical neurophysiologist’s review on the same dataset EEG [62]. Birjandtalab *et al*. used a random forest algorithm to determine which channels contributed the most variation to discrimination of seizure versus non-seizure events [43]. However, their minimal channel reduction, from 23 to 18 channels, provides little difference in clinical application where the patients still wear 18 electrodes. Bhattacharya and colleagues pre-selected 5 out of 23 channels to perform a multivariate analysis of EEG signals [46]. The one channel that displayed the least standard deviation informed the selection of the remaining four channels based on their inter-dependency level and similarity to the first channel. The model’s performance ranged from 0.95 to 0.99, making it comparable to other channel selection methods.

A unique approach by Shah *et al*. focused on domain knowledge to inform the channel selection [48]. They exploited insights on brain hemisphere function, the proximity of a given electrode to other electrodes, electrode position on the scalp, and the region the electrode covered in terms of signal capture. Their eight electrode montage produced the most favourable results, yet the authors affirmed the scarcity of superior techniques that permit electrode reduction whilst maintaining model sensitivity. A rare prospective study by Kjaer *et al*. investigated automated seizure detection in a paediatric population of six patients aged 7–12 years [49]. Using three electrodes, with respective references, they achieved a mean sensitivity of 98.4%, a specificity of 100% and a mean false detection rate of 5.5 per 24/hr. Their study lacked comparisons to a full set of electrodes, but the prospective study design and facilitation of at home device usage made this an exemplar study. Further efforts in the vein of the discussed fundamental works would be invaluable for the seizure detection community.

### 6.1. Limitations of reduced channels

Technological challenges prevail in data collection and hardware design. Stevenson *et al*. [57] found electrode reduction negatively impacted visual interpretation by human experts. They misjudged seizure burden, and seizure annotation was significantly higher when they used 19 rather than eight electrodes. Rubin *et al*. [59] had two epileptologists label cases as seizure or no seizure. When compared to ground truth data, the epileptologists achieved a combined 70% sensitivity and 96% specificity for seizure detection. The reduced arrays were believed to contribute to the inferior sensitivity score. Herta and colleagues compared human expert seizure annotations in ICU EEG recordings with an automated electrode reduction model [4]. They reduced electrodes in a step-wise fashion, calculating sensitivity and specificity for each eliminated electrode. They deemed a minimum of nine electrodes requisite to detect the ictal patterns that the human experts detected.

Channel selection studies highlight the wealth of possibilities in channel selection model combinations. However model combinations, and the interchangeably used terms “electrode” and “channel,” make delineation, comparison and reproducibility of best-in-class models challenging. The impositions described in the literature illustrate the need for further research.

Since EEG is an established and reliable method for capturing and identifying seizure activity, its utility would be amplified if it were evolved beyond the current non-mobile state. A portable device with only a few electrodes would enable long-term recordings, providing rich neurophysiological data for patient-specific data processing. Greater access to data could increase researchers’ proclivity to upgrade existing EEG solutions. Transforming the existing solutions from a diagnosis-only state to a seizure logging and management aid for clinicians and patients will enhance the patient journey. Neurobehavioural and psychiatric comorbidities of epilepsy might be better understood [63,64]. The diagnosis and treatment pathways for epilepsy comorbidities could be optimised [65,66]. Automated seizure detection models could be refined and enhanced by the addition of selective biomarkers. Although the two electrode selection models proposed by our group were encouraging, they signify the difficulty in achieving optimal performance. Despite the first model producing improved results for some, most patients’ performance declined as electrodes were reduced from 19 to 6. The results from the second model were concurrent with the literature, yet the model’s average false alarm score would be unacceptable in a clinical setting. Further prospective studies that elucidate dependable methods for reducing electrodes can pave the way for improving patient care.

## 7. Conclusion

The impact of electrode reduction on both human and computer seizure detection performance is problematic. Deterioration in performance is often attributed to the reduced availability and accuracy of the EEG data. We propose that a resurgence of research into electrode reduction in the clinical setting will support the development of portable, reliable devices, thereby enabling long-term monitoring and enhancing patient quality of life.

## Data Availability

All data produced are available online from the respective dataset owners (Temple University Hospital and the Epilepsy Center, University Hospital of Freiburg).

## Author Contributions

Conceptualization, C.M. and O.K.; methodology, C.M., Y.Y., N.T., A.N. and O.K.; data analysis, C.M., Y.Y., N.T., and O.K.; investigation, C.M., Y.Y. and N.T.; data curation, C.M., Y.Y. and N.T.; Writing-original draft preparation, C.M., N.T., C.W., A.N., and O.K.; Writing-review and editing, C.M., Y.Y., N.T., C.W., A.N., and O.K.; supervision, C.W., A.N., and O.K.; All authors have read and agreed to the published version of the manuscript.

## Acknowledgments

Christina Maher acknowledges the scholarship support from the Nerve Research Foundation. Omid Kavehei acknowledges the partial support provided by The University of Sydney through a SOAR Fellowship and Microsoft’s partial support through a Microsoft AI for Accessibility grant.

